# Interoperability Assessment of Emergency Department Processes Based on Multicriteria Decision-Making Methods

**DOI:** 10.1101/2022.02.21.22271273

**Authors:** Fernanda Wanka Laus, Fábio Pegoraro, Eduardo de Freitas Rocha Loures, Eduardo Alves Portela Santos

**Affiliations:** Industrial and Systems Engineering Program at the Pontifical Catholic University of Paraná (PUCPR), Curitiba (PR), Brazil; Graduate Program in Health Technology (PPGTS) at the Pontifical Catholic University of Paraná (PUCPR), Curitiba (PR), Brazil; Medicine School at the Pontifical Catholic University of Paraná (PUCPR), Curitiba (PR), Brazil

**Keywords:** Emergency Department, Interoperability, Enterprise Interoperability Assessment, Healthcare, AHP, PROMETHEE, MCDA.

## Abstract

It is noticeable that, because of the amount and quality of information exchanged and the criticality of the decisions guided by them, hospitals are considered as some of the most complex organizations in modern society. Evidencing it, emergency departments in hospitals are examples of such complex environments that need perfect integration among systems, people, departments, and data. The processes behind the Emergency Department (ED) routine cause a set of critical and time-dependent decision-making, which must consider several criteria related to organizational and clinical attributes. Based on the literature and worldwide initiatives related to managing complex organizations, an ED can be interpreted through the enterprise interoperability (EI) lens, a concept related to the capability of different systems to work collaboratively within and throughout the organization. Thus, this paper proposes a framework focusing on interoperability assessment in ED processes, where different actors need to interoperate. The proposed framework for Enterprise Interoperability Assessment (EIA) is based on multicriteria decision analysis (MCDA) methods, considering organizational and clinical attributes of the ED. The Analytic Hierarchy Process (AHP) and Preference Ranking Organization Method for Enrichment Evaluation II (PROMETHEE II), in an integrated approach, were elected as the most appropriate multicriteria methods to support, respectively, diagnostic (interoperability assessment) and decisional (interoperability improvements) processes in this background. Last, to validate the framework proposed, a case study was carried out at a stroke unit in a University Hospital (UH) in the south of Brazil. The outcome inferred that the UH achieved the intermediate level of enterprise interoperability in stroke treatment, and Business and Information Technology concerns were found to be the priorities, which deserved greater efforts, to enhance interoperability.

## Introduction

Healthcare (HC) organizations deal with a large number of activities requiring prompt decision-making, which demands assertive knowledge-based and technical skills to be used in short time frames. Such decision-making also requires precise interaction and collaboration between people and departments. Thus, the amount and quality of information exchanged and the criticality of the decisions guided by them make hospitals some of the most complex organizations in modern society (1,2). To ensure the best outcomes for patient treatments, hospitals have to meet the ongoing need of reducing unpredictability, variability, and unexpected outcomes and maximizing the quality and efficiency of their processes [1, 3].

The Emergency Department (ED) reflects this complexity since unassertive decision-making on its part will mean high rates of morbi-mortality. Exemplifying this complexity are two sayings in the field of medicine, “time is brain” and “time is muscle,” to emphasize that the longer one waits to treat a stroke or myocardial infarction, respectively, the more damage is done. The early diagnosis and other measures taken during the initial moments in the ED, which depend on decisions made by different actors and resource flow, are crucial to the prognosis. Likewise, [3–5] are emphatic about addressing the need for perfect integration and collaboration among all players involved in patient care flow for those suffering from acute ischemic stroke.

Furthermore, the importance of optimizing an HC process is highlighted in the COVID-19 era, especially in the ED. Presently, many patients are unwilling to visit a hospital due to fear of being contaminated by the COVID-19 virus. Subsequently, hospitals are facing a worrying scenario wherein patients are being admitted after or almost after the end of the maximum time limit by which they could receive the optimal drug [6, 7]. This time-dependence emphasizes the complexity of clinical processes, impacting the coordination of resource flow and decision-making.

Data and information are essential to assertive decision-making. However, acquiring them in a continuous, sustainable, and reliable manner has proven to be a serious task in the HC field. In a hospital, patient records are often incomplete, especially in an ED where patient care is initiated rapidly. Moreover, there is a dearth of standardization of medical terminology and there are frequent changes in administrative routines [8]. This impaired data exchange is an obstacle to the interoperability of different actors.

Furthermore, information exchange includes communication among departments. [5] state that an integrated and collaborative HC service results in higher patient satisfaction and enhanced cost-effectiveness, while miscommunication among departments is the main cause of unexpected outcomes in health services [9]. From this context blooms the Interoperability concept.

Interoperability is the capability of two or more systems to exchange information and use it as needed, as per its well-known definition [10]. Since it also concerns the practical use of information, it goes beyond health records, being related to the capacity of different entities working together with mutual goals, defining the enterprise interoperability concept [11]. For the Healthcare Information and Management Systems Society, HC interoperability means the capability of different systems, organizations, sectors, and employees to “work together within and across the organization in order to promote the health of individuals and the community” [12].

In short, interoperable systems mean optimal synchronisation between hospital activities, integration of human resources, and maximized efficiency and productivity, whic is likely associated with increased operational benefits [13]. Faced with this scenario, it is inferred that social-technical and interoperable HC systems are mandatory in HC, driving the understanding of the ED’s treatment processes from the enterprise interoperability perspective [14]. From this approach, an organization could be submitted to an Enterprise Interoperability Assessment (EIA), a process that allows for the identification of its strengths and weaknesses and, thus, prioritize actions to improve its interoperability status [11]. To illustrate the advantage of this system of assessment, the West Health Institute estimated that the US HC sector lost approximately 30 billion dollars due to interoperability failures [15], which could have been minimized through EIAs.

Pointing out the necessity of optimizing HC treatment processes as well, the “Joint Commission on Accreditation of Healthcare Organizations” in the US reported that the main causes of sentinel events occurring in the US HC sector in 2014 were related to cognitive failures. Thus, hospitals should focus on minimizing human variability to mitigate risks [16].

Once the importance of information exchange and integration of different departments in a hospital is clear, the enterprise interoperability approach can elucidate improvements in some specific clinical processes. This paper considered the Analytic Hierarchy Process (AHP) a suitable Multicriteria Decision Analysis (MCDA) method to assess enterprise interoperability, once AHP is capable of organizing knowledge by arranging the elements in a hierarchical structure and enabling multiple evaluations with many conflicting particularities [17, 18]. Subsequently, the diagnostic approach is complemented by visualizing ways to improve the current interoperability, a prognostic approach, which is well supported by the Preference Ranking Organization Method for Enrichment Evaluation II (PROMETHEE II), another MCDA method. The PROMETHEE II is an outranking method; it makes explicit whether, through ranking the alternatives, an attribute is already delivering sufficient performance or needs improvements [18, 19]. The use of a hybrid MCDA approach offers more robust results than isolated MCDA methods [20].

Following this rationale, this paper proposes a framework for EIA specifically designed for ED processes that is composed of two stages: the first one is the interoperability assessment (i.e., it elucidates which interoperability level the ED is in); the second stage must guide hospitals, based on the interoperability diagnostics, in prioritizing the efforts to improve their interoperability potential. This framework was applied in the stroke unit of a University Hospital (UH) in the south of Brazil. A stroke unit, an ED focused on stroke care, is a department designated for patients with acute stroke, including continuous monitoring of vital parameters as well as specialized staff.

Thus, this article is structured as follows. The second section presents a literature review on enterprise interoperability concepts and the usage of MCDA methods relating them to the HC field. The third section proposes and formulates the framework for EIA. The fourth section presents the implementation of the framework in the University Hospital and discusses the results obtained. Conclusions and final considerations are discussed in final section.

## Literature Review

This Section presents the theoretical concepts that underpin this interdisciplinary research based on scientific literature. Enterprise interoperability and its fundaments are conceptualized, and the MCDA methods are detailed.

### Enterprise Interoperability in Healthcare

Interoperability was first defined by the Institute of Electrical and Electronics Engineers (IEEE) as “the capability of two or more systems to communicate and exchange information with each other and use this information” [10]. Of late, this concept has received a broader meaning, so that enterprise interoperability may be understood as the capability of inter and intraorganizational systems to work together with mutual objectives [11]. [21] highlight that systems should be understood from a generic perspective, wherein a system could be a stakeholder, an enterprise, a department, an information system, or a resource. This understanding enables one to comprehend the HC domain, populated by different kinds of systems, under the interoperability concerns.

Two classical models in enterprise interoperability, ATHENA Interoperability Framework (AIF) and Framework of Enterprise Interoperability (FEI), have introduced the concept of Interoperability concerns, considering that enterprise interoperability could be analyzed from different standpoints. Both AIF and FEI consider four Interoperability concerns: business, process, service, and data [21, 22].

These concerns do not contemplate the human element but focus on the structural and organizational ones. However, HC treatment outcomes tightly depend on human variants facing the need for multidisciplinarity: the variability of each case, the individuality of each patient, and the physicians’ individual experiences influence medical decisions [2]. Following this, [23] proposed another model, which divides interoperability into human, non-human, and heterogeneous systems, and then evaluates overall enterprise interoperability. Thus, these authors highlighted human systems for the academic community inclined toward interoperability.

Further, [24], in a literature review, identified that none of the existing models with maturity levels at par with those of interoperability could verify the existing weaknesses that impair interoperability. Therefore, they proposed other concerns that would best fit human-dependent contexts: (i) business (strategic aspects such as culture, vision, values, and strategy policies); (ii) process management (related to working methods); (iii) human resources (skills, competencies, roles, collaboration, and social culture); (iv) policies and procedures (normative aspects such as standards and guidelines); (v) information technologies (interconnecting applications, data, and communication); and (vi) semantics (the understanding of different terminologies).

Healthcare interoperability frameworks in the literature are termed as E-Health, Health Information Systems (HIS), and Personal Health Systems (PHS) [25]. However, these preexistingframeworks only focus on health records and hospitals strategy, not considering attributes of clinical processes. On the other hand, the framework proposed by this paper aims to analyze the enterprise interoperability achieved by critical clinical treatment processes, such as those of the emergency department.

### MCDA

The MCDA (multicriteria decision analysis) is a discipline that evaluates multiple conflicting criteria in decision-making, facilitating the choice of a solution in scenarios with several qualitative and quantitative variables, useful for benefit-risk analysis. As the volume and complexity of health data, information, and knowledge is growing, the MCDA has become invested with more potential in optimizing decision-making at any level of HC [26–28]. Its popularity in HC is explained by the large variety of attributes that exist in such a context [29]. The bibliometric analysis conducted by [29], which reviewed 66 papers on the MCDA in the HC field, revealed that 39% of them used the MCDA aiming for the “diagnosis and treatment” subarea, and the second and the third subareas more prevalent were “priority setting” and “health technology assessment,” both representing 12% each. [30] reasoned that MCDA could be applied to any field of emergency medicine. Regarding public health, [28] affirm that, recently, MCDAs have been more utilized because, conducting a systematic scientific empirical assessment designed to evaluate medical technology, they allow government managers and agencies to support appropriate decisions on healthcare.

### AHP

[31] pointed out that 26% of all papers published on the AHP, until the year of their study, contained information on the employment of the AHP in managerial evaluation models (such as those pertaining to quality and performance levels). The other papers reviewed by them applied AHP on selection, benefit-cost, prioritization, resource allocation, and medicine. Cases of strategic plan-related choices, including ones related to a HC processess, consist of a set of AHP solutions presented in [32]. The AHP has been then widely used for facilitating HC decisions [27].

A recent paper described how AHP developed over time, providing a historical perspective from 1979 to 2017 [33]. This review found that in the first years (1979-1990) AHP was majority applied on mathematics, eventhough other fields, such as business and managements, health and social issues, economics, and computer science, were becoming potential AHP users. Then, from 1990 to 2017, AHP has been widely applied in the health sector, frequently proposed as a method to evaluate different medical treatment strategies. This study also concluded the application of AHP integrated with others MCDA methods was emphasized amid 7639 papers published between 2002 and 2017 [33].

The AHP allows to model some problem in a hierarchical structure, deploying it downward from a goal, subsequently branching it into criteria and subcriteria and, in the end, leading it to solution-based alternatives. The number of layers and elements in each level varies according to the problem analyzed. Nonetheless, the model must be complete enough to facilitate an understanding of the situation and simple enough to facilitate being sensitive to small changes. The use of the AHP can be summarized as three aspects: break down of the problem into a hierarchical structure, pairwise judgments, and synthesis of priorities in order to obtain a prioritization of the alternatives involved, indicating the best alternative [34].

The use of AHP has received attention in EIA-oriented applications [35–38]. The EIA caracterizes of the goal of the AHP, while the attributes involved in the domain must be in the layers of the criteria and subcriteria and, after all, the AHP alternatives indicate the level of interoperability achieved. The corresponding AHP model is summarized in Fig 1. Pairwise comparisons must be made between each element of the problem according to the Saaty Scale (ranging from 1 to 9 [Table 1]). Several people involved in the process must judge these weights [34]. The mathematical foundations behind the AHP are summarized below.

**Fig 1.**
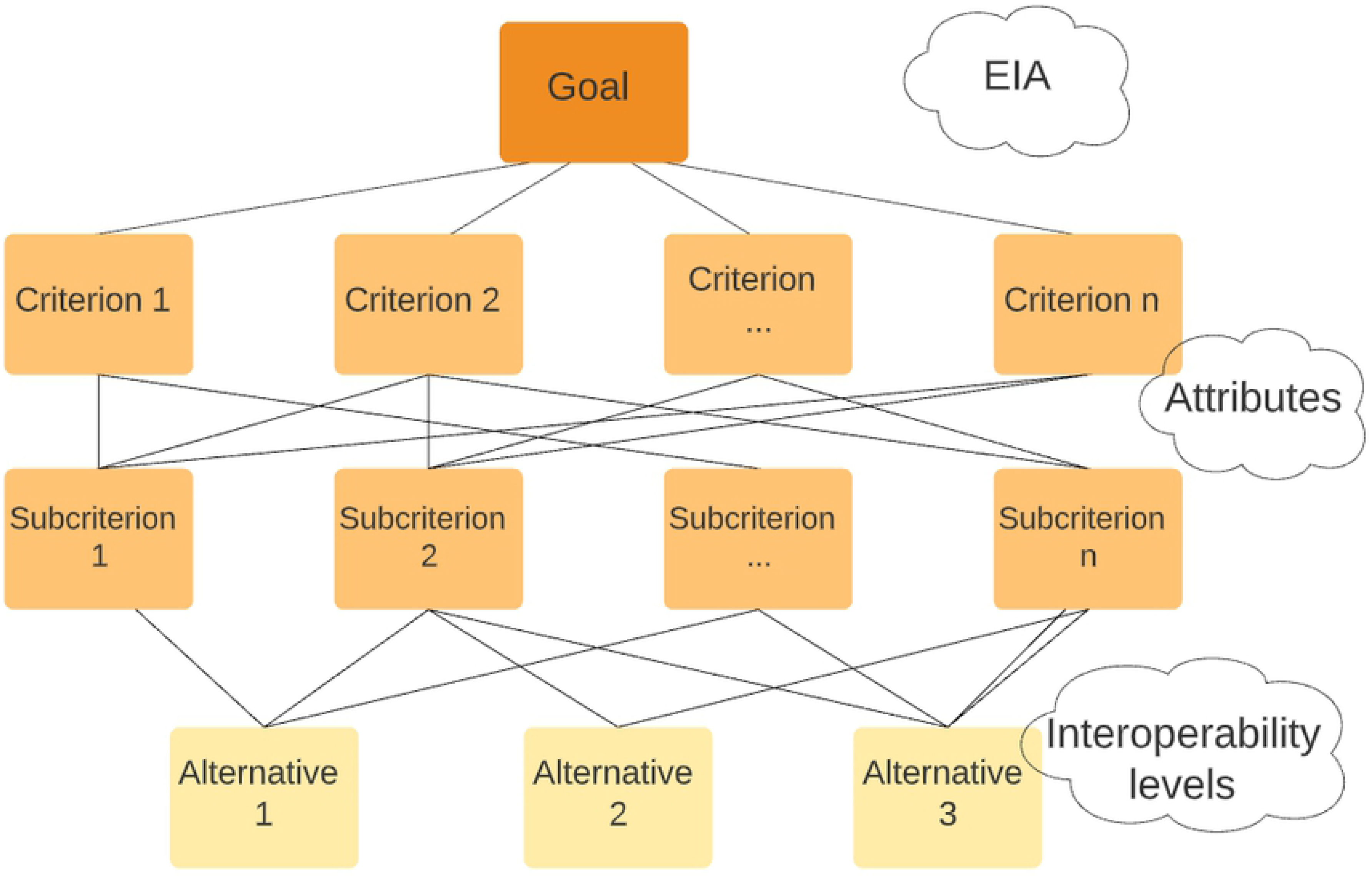
EIA structured by the AHP method.

**Table 1.** Saaty Scale.

| Definition | Numerical Rating | Reciprocal |
| --- | --- | --- |
| Both equally important to the objective | 1 | 1 |
| Moderate importance | 3 | 1/3 |
| Strong importance | 5 | 1/5 |
| Very strong importance | 7 | 1/7 |
| Extreme importance | 9 | 1/9 |
| Intermediate values between two adjacent values | 2, 4, 6, 8 | 1/2, 1/4, 1/6, 1/8 |

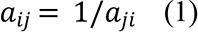

- Pairwise comparisons between two elements according to the Saaty Scale, as follow.
- Construction of reciprocal matrix *A*_*ij*_ with (*n* (*n* ― 1)) elements.
- Normalized reciprocal matrix *A*′_*ij*_: divide each element by the sum of the elements of that column, resulting in the column-sum reduced to 1.
- Eigen vector *W*: priority weighting of each element calculated according to Equation 2, where *W*is the eigen vector and λ is the eigen value.

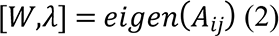

- Normalized eigen vector *W*′: is the priority vector. Each element is multiplied by 100% to obtain the ranking. In Equation 3, *x*% means the prioritization of *x* in the final ranking.

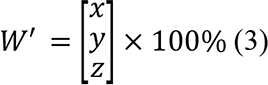

- The consistency of the ranking must be checked by calculating the Consistency Ratio (*CR*) according to Eq. 4, where *RI* is the Random Index. There are standard values for *RI*, as illustrated in Table 2.

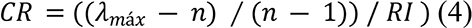

- If *CR* < 10%, then the judgments are consistent, and the results are valid. If not, the judgments must be reviewed.

### PROMETHEE II

PROMETHEE is a set of MCDAs proposed by Brans & Mareschal in 1982. The earliest research projects using them in a HC context were published in 2002, and they studied the prioritization of a given resource across hospital departments [39, 40]. Public health organizations should be encouraged to to adapt, use and share PROMETHEE as an effective approach to enable the integration of multiple perspectives in the control of public healtht issues [26]. The PROMETHEE methods also use pairwise comparisons and prioritizations. However, they consider the performance of attributes. The comparisons result in over-classifications representing the performance of the alternatives with respect to a specific criterion/attribute [19].

**Table 2.** Random Index values according to *n*.

| **n** | 1 | 2 | 3 | 4 | 5 | 6 | 7 | 8 | 9 | 10 |
| --- | --- | --- | --- | --- | --- | --- | --- | --- | --- | --- |
| **RI** | 0.00 | 0.00 | 0.58 | 0.90 | 1.12 | 1.24 | 1.32 | 1.41 | 1.45 | 1.49 |

Among the PROMETHEE methods, the PROMETHEE II has been employed in this study. It admits incomparability between the alternatives with the classification “indifference,” unlike other MCDAs. There are three classifications for comparisons: preference, indifference, and incomparability. PROMETHEE II also considers a net flow (balance between negative and positive flows), a limitation of PROMETHEE I, where the ordering does not compare conflicting actions [19]. The final selection of the PROMETHEE II ranking is based on each alternative’s positive and negative preference flows. A positive preference flow indicates the extent to which an alternative outperforms the others, while a negative preference flow indicates the extent to which an alternative is outperformed by the others. Thus, PROMETHEE II provides a performance ranking for each of its elements, ideal to understand whether a criterion already has exhibited enough performance and to what extent or it needs to be improved and to what extent. The mathematical foundations are summarized as follows [19].

- Definition of alternatives and criteria: calculate the deviations between the evaluations of the alternatives, according to Equation 5, where {*g*_1_, *g*_2_,…, *g*_*n*_} = set of evaluation criteria.

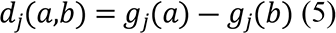

- Information between criteria: the weightings of relative importance of the different criteria must be normalized, according to Equation 6.

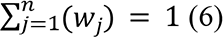

- Information within the criteria: calculate the Preference Function, according to Equation 7, which means that the larger the deviation, the larger the preference.

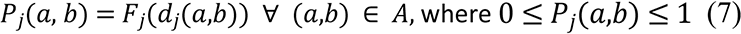

- In case a criterion has to be maximized, this property holds Equation 8, and if needs to be minimized, the preference function should be reversed.

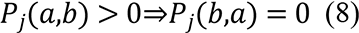

- Six preference functions have been proposed: usual criterion, U-shape criterion, V-shape criterion, level criterion, V-shape with indifference criterion, and Gaussian criterion.
- Aggregated preference indices are calculated. The result of Equation 9 shows how *a* is preferred to *b* over all the criteria. The result of Equation 10 shows how *b* is preferred to *a* over all the criteria.

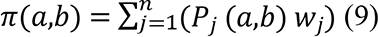

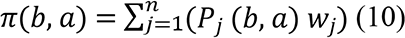

- Outranking flows: each alternative is facing (*n* ― 1) other alternatives in *A*. The positive and the negative outranking flows are calculated by Equations 11 and 12.

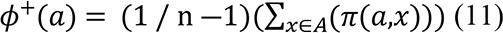

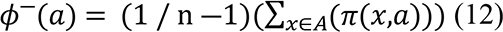

- The final ranking is obtained by using Equation 13.

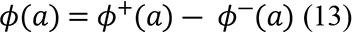

### Proposal of an Enterprise Interoperability Assessment **framework**

Systems are intrinsically linked to the enterprise interoperability concept. In sum, a system is a set in which a limit can be established, and the elements within it are the attributes that characterize and differentiate it from others. Thus, attributes are the elements that define the structure of a class that allow the qualification and relation of confidence and commitment between interoperable systems [25]. Following this rationale, it is necessary to identify the attributes that define critical HC processes.

Personal Health Systems (PHS), an interoperability framework, considers human aspects in addition to technical ones, in line with the interoperability concept propounded by [23] and later reinforced by [24]. The PHS is divided into two structures: execution and technical aspects; legal and organizational aspects [25]. Based on this distinction, HC attributes could be divided into organizational aspects related to legal and organizational matters and the clinical aspects referring to treatment execution and techniques. The attributes existing in a HC process directly interfere with its clinical outcomes as well as face interference by enterprise interoperability.

In this sense, this research proposes a framework capable of diagnosing the interoperation capacity and provide guidance on how to improve it (i.e., a set of actions suggested to improve interoperability to optimize clinical outcomes). The framework is subsequently based on two stages: (S1) diagnosis of the current scenario of interoperability (EIA); (S2) establishment of a decisional basis, which prioritizes actions for improvement. The stages S1 and S2 and their relationship with clinical outcomes and attributes comprise the framework shown in Fig 2.

**Fig 2.**
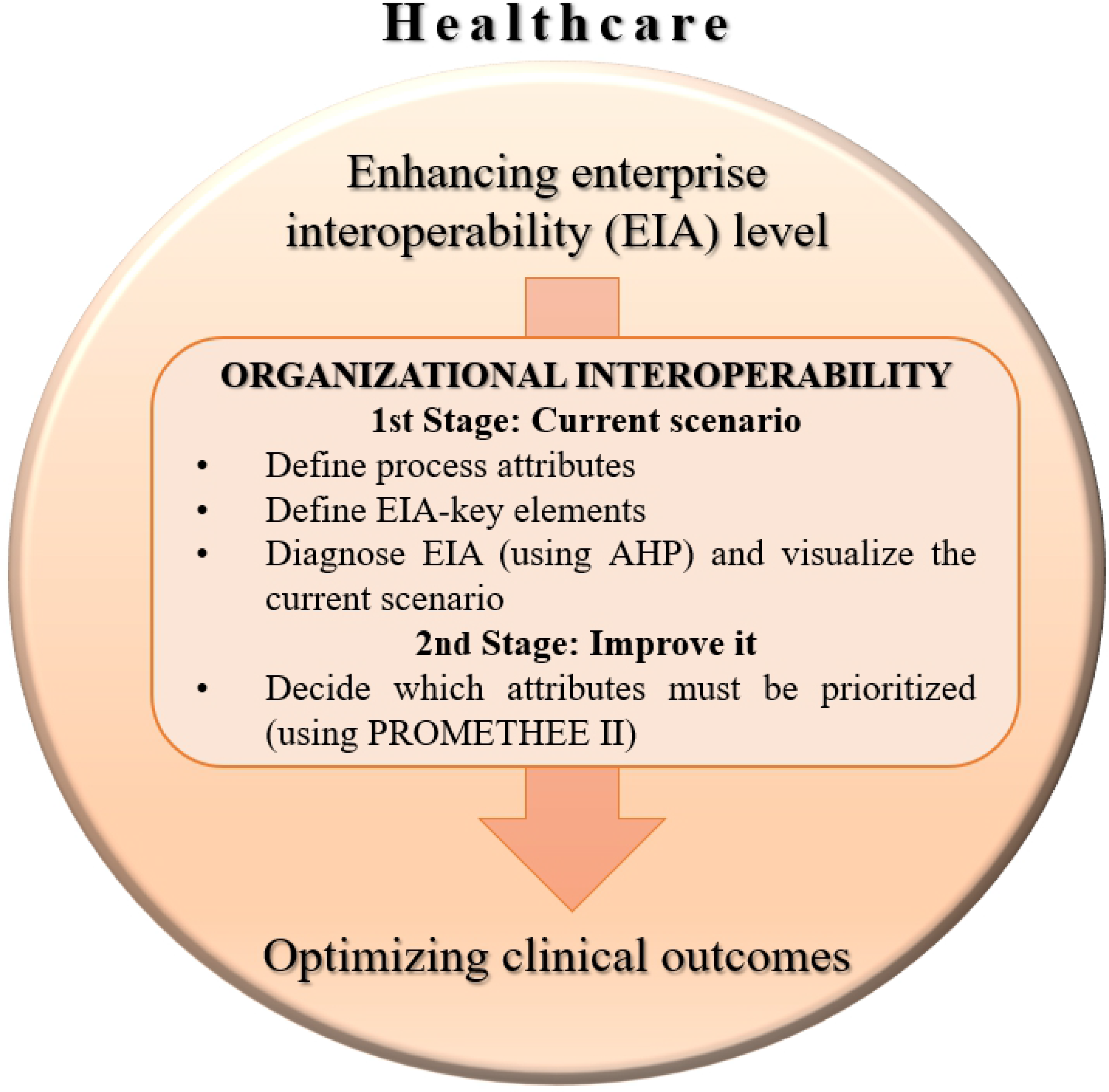
Illustration of the Framework, containing its two main stages.

The definition of the key elements used in this EIA was a prerequisite for the first stage (S1). The organizational and clinical attributes, interoperability concerns, and interoperability levels had to be postulated. The organizational attributes (OAs) were extracted from the literature, while the identification of the clinical ones, in addition to the literature, was contributed to by physicians working in the public and private sectors. The framework and its stages could be performed after defining these key elements.

Further, both stages employed MCDA methods. The first one, (S1), used AHP, which performed the EIA, pointing out a level of interoperability corresponding to the “as is.” In sequence, the second stage, (S2), utilized PROMETHEE II, based on S1, to weight the criteria, and its outcome was a ranking of interoperability concerns, which prioritized the attributes that led to a more advanced level of interoperability. Thus, S2 helped decide which attributes must be prioritized in terms of efforts for improvement.

Hybrid MCDA approaches can offer more robust results than isolated MCDA methods [20]. [41] listed that, until 2010, 48 papers used the PROMETHEE associated with other MCDA methods, specially the AHP. The employment of these two methods in a complementary way is based purely on their definitions. The AHP enables a top-down analysis, and it is used to define and obtain the weighting of importance for criteria and alternatives, based on expert opinion and literature research. The PROMETHEE II uses the data from the first method to allow for a final ranking of alternatives, providing the prioritization of actions for improvement [42, 43]. This paper is novel in that it combines the hybrid MCDA approach and EIA, focusing on improvements of HC processes since the framework was applied at a stroke center. Section 4 describes the case study based on this framework.

### 1st Stage: Diagnostic basis – Enterprise Interoperability Assessment

#### Definitions of the key elements

This subsection postulates the key elements of the EIA, i.e., the attributes, interoperability concerns, and levels of interoperability, once they are its prerequisites [25]. The OAs, related to hospital management, were gathered based on those proposed by (17). Table 3 lists these attributes.

**Table 3.** Organizational attributes related to hospital management.

| Abbreviation | Organizational Attributes |
| --- | --- |
| OA 1 | Reception / Registration / Triage |
| OA 2 | Electronic medical record |
| OA 3 | Control of physical files and records |
| OA 4 | Relationship with external entities |
| OA 5 | Intersectoral communication coordination |
| OA 6 | Technological resources to support services |
| OA 7 | Intraorganizational communication via system |
| OA 8 | Simple and easily accessible information systems |
| OA 9 | Technical and functional system support |
| OA 10 | Electronic service protocols common to the whole system |
| OA 11 | Protocols and procedures updated with current guidelines |
| OA 12 | Compliance between practice and standards/protocols |
| OA 13 | Coordination of clinical information flow |
| OA 14 | Means of communication between hospital and patients |
| OA 15 | Information available to the patient |
| OA 16 | Research and development of new technologies and treatments |

In parallel, the clinical attributes (CA) concerning the processes of the ED related to the treatment of patients with the most recurrent and critical diseases were listed. The essential elements common to the most critical clinical conditions were identified based on elemental bibliography of internal medicine [44] and, subsequently, confirmed by interviews with experts (intensivists, neurologists, and internists were interviewed). Table 4 lists the CAs. It should be noted that this is a summary containing key elements of management, with no interest in specific clinical details.

**Table 4.** Clinical attributes within the scope of stroke treatment.

| Abbreviation | Clinical Attributes |
| --- | --- |
| CA 1 | Initial measurement of vital signs |
| CA 2 | Glasgow Coma Scale on admission |
| CA 3 | Specific scale evaluation |
| CA 4 | Imaging examination reports by a specialist |
| CA 5 | Access to patient's prior history |
| CA 6 | Differential diagnosis |
| CA 7 | Neurological status |
| CA 8 | Constant monitoring of vital signs |
| CA 9 | Administering drugs for clinical stabilization |
| CA 10 | Quick access to imaging examinations |
| CA 11 | Quick access to investigation examinations (e.g., ECG, laboratories) |
| CA 12 | Establishing the exact time of symptom onset |
| CA 13 | Administering the specific drug needed |
| CA 14 | Fitting the patient's condition into the set of inclusion or exclusion criteria for specific treatment |

For instance, all cases involve the reading of a list of clinical characteristics which physicians must observe to judge whether the patient should be administered the first choicedrug. When the “therapeutic window,” a period of time, is exceeded, it disables the administration of the specific drug. Other clinical characteristics that should be evaluated are comorbidities, drugs in use, and previous surgeries, evidencing the need for robust databases to have quick access to such information. Other CAs concern labs and imaging examinations, their disponibility and accuracy, and whether their reports are formulated by specialists.

Evaluating the Glasgow Coma Scale and other specific scales, measuring and monitoring the vital signs, monitoring the neurological status, and judging the differential diagnosis are also common attributes in the ED. Once the need for interoperation among different systems in a short time interval was clarified, the CAs within the scope of stroke treatment were listed as follows (Table 4).

Regarding the concepts pertaining to EIA, it was important to define the most suitable interoperability concerns and levels. This framework took into consideration the enterprise interoperability concerns proposed by (24), namely, business, policies and procedure, process management, human resources, semantics, and information technology. Table 5 summarizes them. Next, the interoperability levels were adapted from [45], who studied EIA in the HC domain. As a result, three levels were established, as described in Table 6.

**Table 5.**
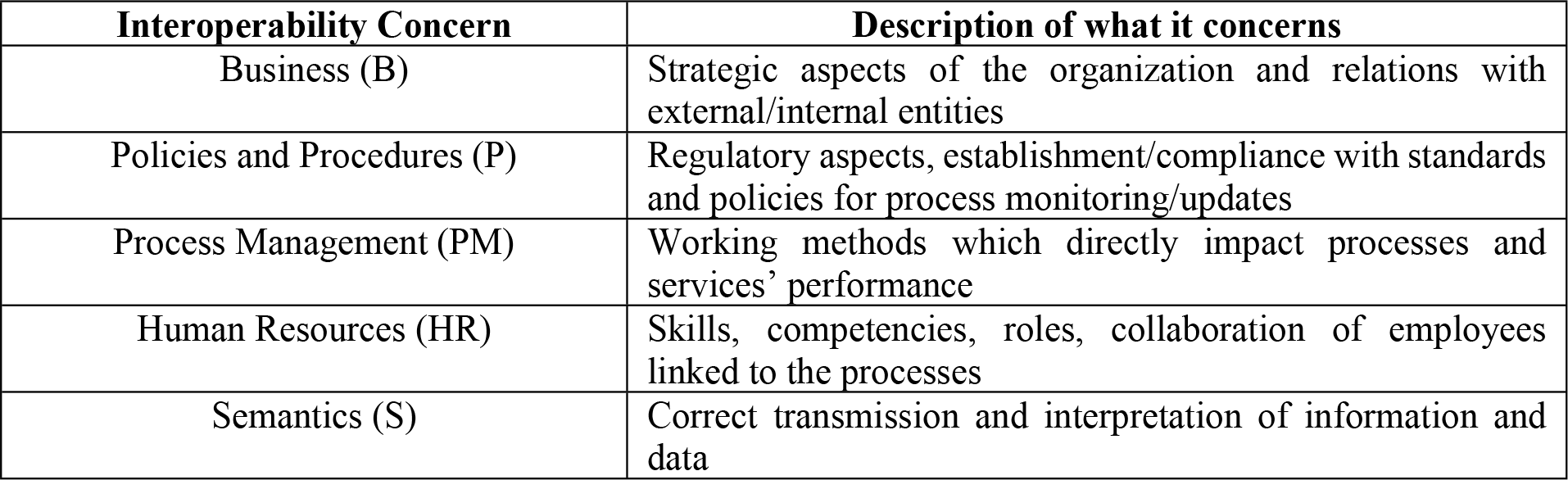
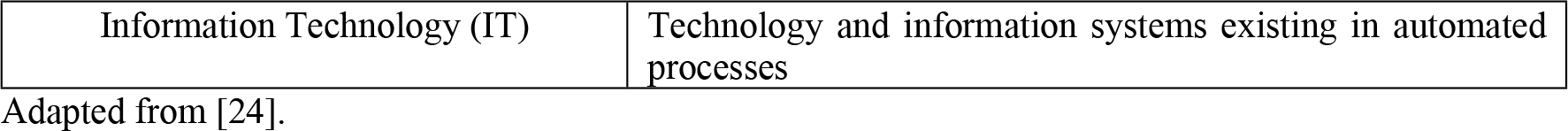
Interoperability concerns and their descriptions.

**Table 6.** Interoperability levels and their description.

| Interoperability Levels | Description |
| --- | --- |
| Basic Level | The institution has no or insufficient capacity to address clinical emergencies, being unable to support the whole patient care. There is a variation in its processes and practices, and it depends on analogical systems. |
| Intermediate Level | The institution admits and manages clinical emergencies, but is unable to dispense the best existing options of treatment in the whole patient care. It seeks reducing variability of its processes and increasing collaboration and integration and has complex systems and processes governed by a central body. |
| Advanced Level | An institution at this level works with extremely developed policies and resources. Moreover, it is constantly innovating and seeking partnerships. Its processes are effective and concrete, designed with the purpose of achieving satisfactory results in the treatment of patients admitted in emergencies. |

Adapted from [45].

#### AHP Design

The AHP’s goal is represented by the EIA itself; the criteria are the interoperability concerns, the subcriteria are the organizational and clinical attributes, and the alternatives (the solution) are the interoperability levels. In order to allocate each element (attributes, concerns, and levels) in the AHP’s hierarchical structure and make possible the pairwise judgments, matrices and a questionnaire were used. Quality Function Deployment (QFD)-inspired matrices were employed to establish the relationships between the interoperability elements, making them explicit, and, thereby, generating entries for the AHP [17,31,37].

The pairwise comparisons between elements within the same AHP layer were made in the upper part of the matrices, following the Saaty Scale (Table 1). Meanwhile, the relationships between elements of different AHP layers, that considered the scale in Table 7, were in the lower part.

**Table 7.** Relationship scale used in the upper part of the matrices.

| Relationship meaning | Very strong | Medium | Weak | No relationship |
| --- | --- | --- | --- | --- |
| Qualitative value | 9 | 6 | 3 | 1 |

The first matrix analyzed the interoperability concerns and OAs. The importance of the concerns for the HC environment was weighted, resulting in Fig 3-A. On the other hand, Fig 3-B contains the pairwise comparisons of the OAs among themselves, and Fig 3-C has the correlations between OAs and interoperability concerns. A specialist with the know-how to judge OAs from the interoperability concern helped with both judgements (Fig 3-B and 3-C).

**Fig 3.**
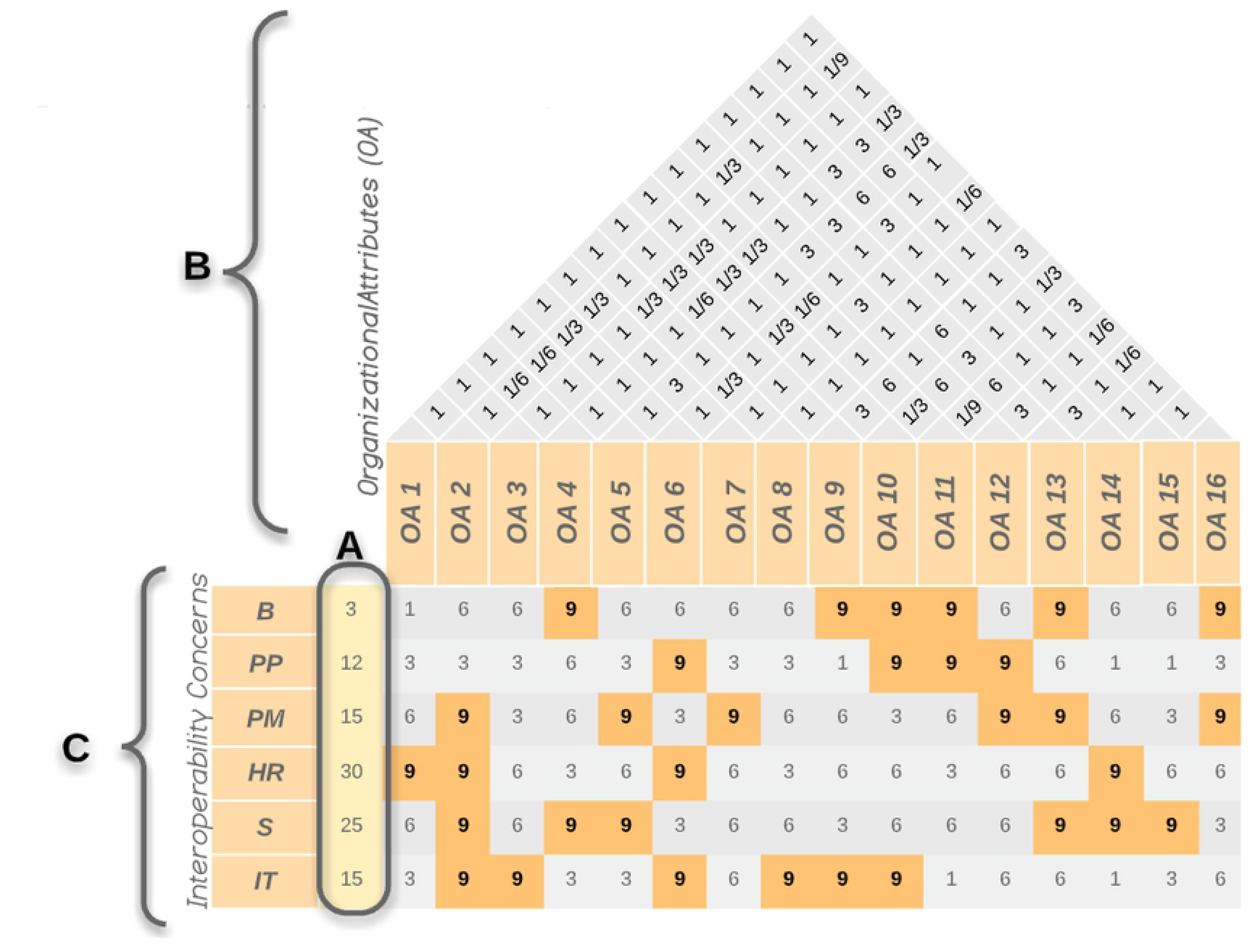
Matrix correlating Interoperability concerns with organizational attributes and comparing organizational attributes to each other. (A) The importance of each concern. It is mandatory to structure the AHP design the (B) pairwise comparisons and (C) correlations between different layers.

The second matrix had the OAs in its rows and CAs in its columns; thus, it gave way to pairwise judgements between CAs (Fig 4-A) and relationship judgements between clinical and organizational attributes (Fig 4-B).

**Fig 4.**
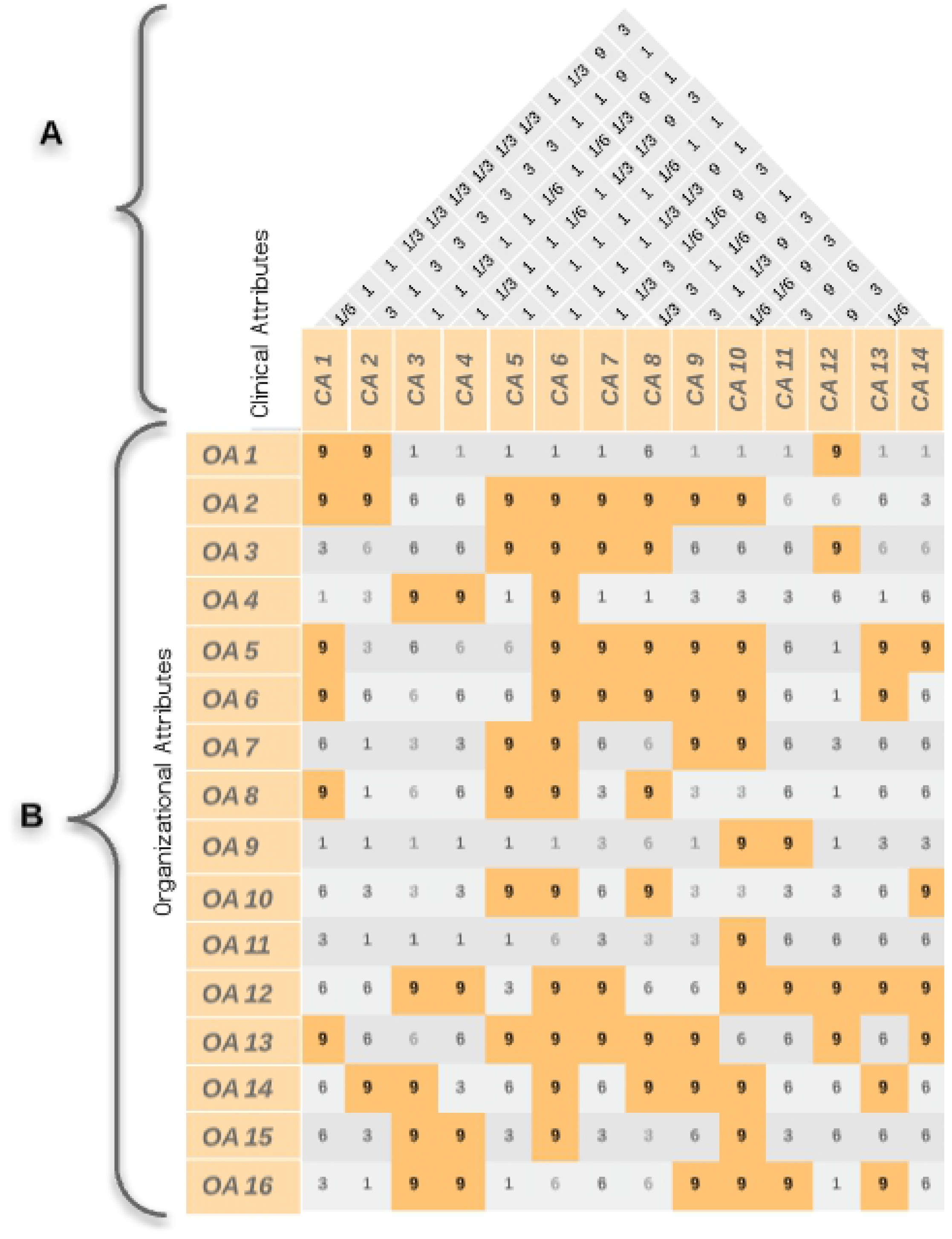
Matrix correlating clinical attributes to organizational attributes and comparing clinical attributes to each other. (A) Pairwise comparisons. (B) Correlations between different layers.

When incorporating the elements into the AHP hierarchical structure, only “very strong” (weight 9) relationships were considered. Tables 8 and 9 list these correlations.

**Table 8.** List of relations between healthcare interoperability concerns and organizational attributes.

| Healthcare Interoperability concerns | Organizational attributes (OA) |
| --- | --- |
| Business (B) | OA 4; OA 9; OA 10; OA 11; OA 13; OA 16 |
| Policies and Procedures (P) | OA 6; OA 10; OA 11; OA 12 |
| Process Management (PM) | OA 2; OA 5; OA 7; OA 12; OA 13; OA 16 |
| Human Resources (HR) | OA 1; OA 2; OA 6; OA 14 |
| Semantics (S) | OA 2; OA 4; OA 5; OA 13; OA 14; OA 15 |
| Information Technology (IT) | AO 2; OA 3; OA 6; OA 8; OA 9; OA 10 |

**Table 9.**
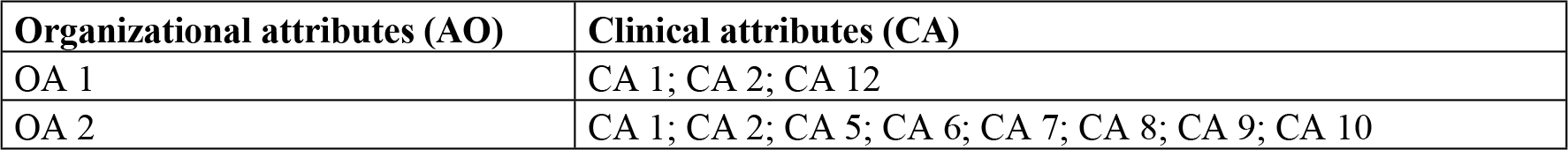
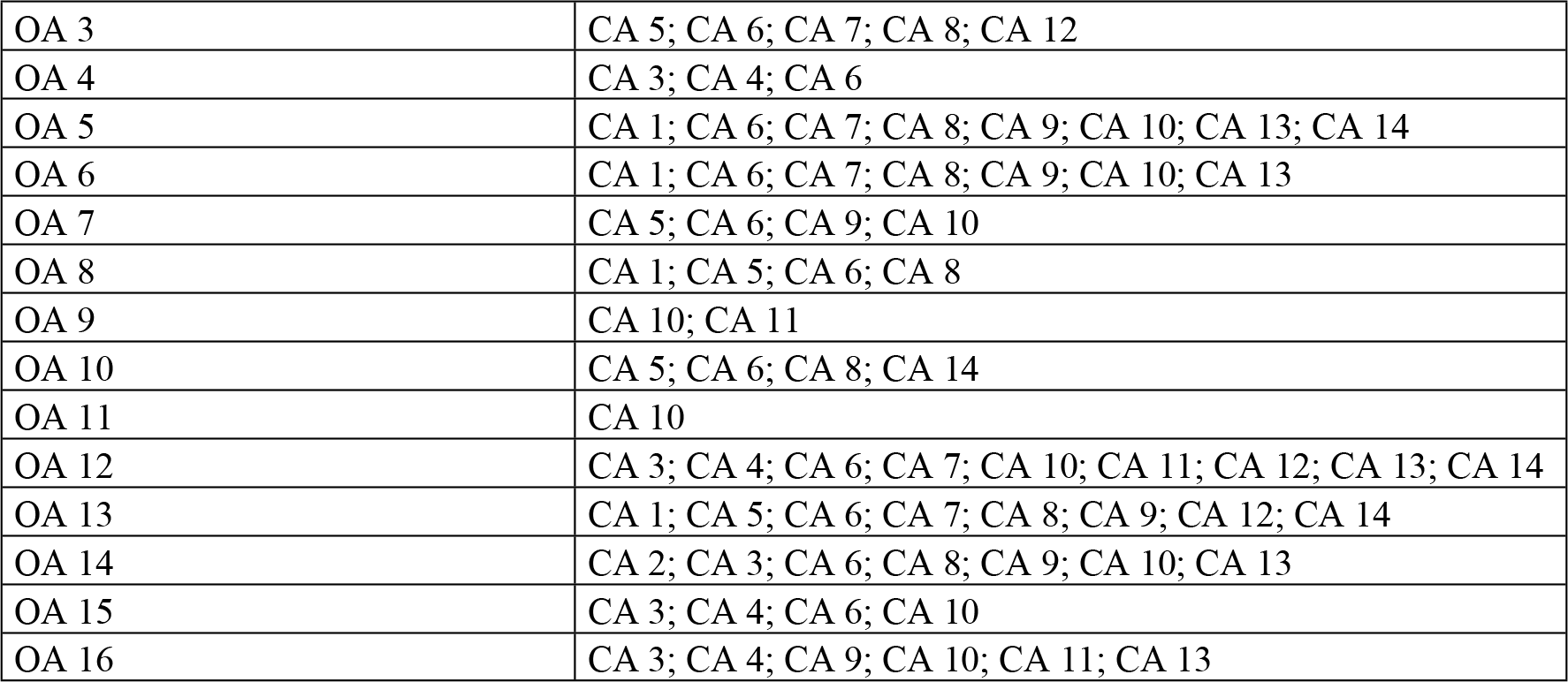
Relations between organizational and clinical attributes in healthcare interoperability.

| Organizational attributes (AO) | Clinical attributes (CA) |
| --- | --- |
| OA 1 | CA 1; CA 2; CA 12 |
| OA 2 | CA 1; CA 2; CA 5; CA 6; CA 7; CA 8; CA 9; CA 10 |
| OA 3 | CA 5; CA 6; CA 7; CA 8; CA 12 |
| OA 4 | CA 3; CA 4; CA 6 |
| OA 5 | CA 1; CA 6; CA 7; CA 8; CA 9; CA 10; CA 13; CA 14 |
| OA 6 | CA 1; CA 6; CA 7; CA 8; CA 9; CA 10; CA 13 |
| OA 7 | CA 5; CA 6; CA 9; CA 10 |
| OA 8 | CA 1; CA 5; CA 6; CA 8 |
| OA 9 | CA 10; CA 11 |
| OA 10 | CA 5; CA 6; CA 8; CA 14 |
| OA 11 | CA 10 |
| OA 12 | CA 3; CA 4; CA 6; CA 7; CA 10; CA 11; CA 12; CA 13; CA 14 |
| OA 13 | CA 1; CA 5; CA 6; CA 7; CA 8; CA 9; CA 12; CA 14 |
| OA 14 | CA 2; CA 3; CA 6; CA 8; CA 9; CA 10; CA 13 |
| OA 15 | CA 3; CA 4; CA 6; CA 10 |
| OA 16 | CA 3; CA 4; CA 9; CA 10; CA 11; CA 13 |

These judgments and correlations, obtained through the literature and expertise of specialists, helped build the two matrices and they, in turn, enabled the AHP design. This is part of the framework construction that is to be replicated in all clinical emergency processes, while the EIA diagnoses the enterprise interoperability level of a specific hospital.

In sequence, the design of the AHP allows for the EIA to proceed. It is necessary that an HC professional, directly involved in the assessment process, evaluates the CAs. To facilitate this assessment by professionals who are not used to interoperability semantics, a questionnaire is employed, whose answers must be translated into the pairwise judgements of the AHP (46). In this way, the questionnaire respondent must evaluate their satisfaction regarding the performance of the CAs and answers should be translated into interoperability levels to best suit the AHP design in such a way that attributes with “unsatisfactory,” “partially satisfactory,” and “very satisfactory” performances fit, respectively, into Basic, Intermediate, and Advanced levels of interoperability. After diagnosing the level of enterprise interoperability in relation to the domain under study, the 1^st^ Stage was concluded.

### 2nd Stage: Decisional basis – Enterprise Interoperability Assessment

Therefore, the 2^nd^ Stage (S2) encompasses a decisional basis, in order to indicate the way toward leveraging the enterprise interoperability. The results of S2 indicate which HC interoperability concerns should be prioritized to increase the interoperability level and how.

#### The PROMETHEE II Design

The inputs for the PROMETHEE II are the weightings of the OAs provided by the AHP (which calculates weightings for each of the OAs, as well as for each of the interoperability concerns that the attributes are related to).

A global weighting (*w*_*OA*_) is established for each organizational attribute following Equation 14.

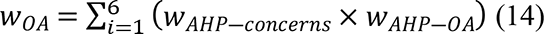

Thus, the global weighting of an OA is defined by the sum of all products of the weightings of the concerns and the OA linked to them Where *W*_*AHP*—*concerns*_ means the AHP weightings of the concerns and *W*_*AHP*—*OA*_ means the AHP weightings of the OA.

While a high-value weighting has a positive inference in the AHP, in the PROMETHEE II, on the other hand, the higher the weighting, the weaker the attribute, i.e., the weightings of attributes have opposite inferences in the AHP and PROMETHEE II and only have equivalent mathematical inferences if inverted. For instance, if the highest AHP weighting (already at a satisfactory interoperability level) is also allocated to the PROMETHEE II as the highest one, an attribute that has already been performing well would be mistakenly interpreted as one that deserves improvement and vice-versa. Due to it, the weightings from the AHP should be reversed in order to identify which interoperability concerns deserve greater efforts.

Complying with the PROMETHEE II’s terminology, interoperability concerns were defined as “actions” and the OAs as “criteria.” Since all the criteria were ascending functions, they were designated as maximum. The best suitable preference functions were chosen according to the matrix structure and performance of criteria (OA). A qualitative impact scale was employed to evaluate the influence that each OA had over each of the interoperability concerns.

For each concern, two indices must be calculated: the positive flow *ϕ*^+^and the negative flow *ϕ*^―^. The symbol *ϕ*^+^ expresses how much a concern needs to overcome the others by, while the symbol *ϕ*^―^ indicates how much its performance is already sufficient by to deliver the desired level of interoperability [47]. A sensitivity analysis function enables an understanding of how to make the improvements, since it relates the concerns’ changing behavior to changes in OA prioritization. As a result, a sensitivity analysis is relevant to indicate which OA should be prioritized.

#### EIA Framework – Case study

The framework proposed by this paper was applied in a UH. This UH is one of the largest hospitals in the south of Brazil and has a high volume of patients admitted in the ED, including some with ischemic stroke symptoms. Even though the UH has a residency program in neurology and neurosurgery, it has acknowledged that its performance in stroke patient management can be improved further.

The most efficient therapeutic procedure to avoid increasing the infarcted brain area is the infusion of intravenous thrombolytic recombinant tissue plasminogen activator (rtPA). However, although rtPA doubles the chance of recovering without sequelae, it should be prescribed with great caution, respecting several indications and contraindications, due to the risk of causing cerebral hemorrhage as a common adverse effect. Therefore, a thorough decision-making process for establishing the patient’s eligibility for such conduct is essential. In addition, the clinical benefit of rtPA is only achieved when infusion occurs within 4.5 hours of the onset of the symptoms, as predicted by [48] and, currently, widely accepted in medical practice. The time elapsed before onset of treatment, agility in diagnosis, prioritization in the patient’s transportation, communication among health professionals, and access to neuroimaging are some of the factors that strongly interfere with morbi-mortality rates.

The complexity of this treatment leads to the need for integrated teams, resources, and infrastructure such that the risk of sequelae and death are minimized [(3–5]. Since perfectly integrated systems contribute to better clinical conduct, the optimization of the process is directly related to the level of interoperability of the hospital entity.

### 1st Stage: Development and the AHP structure

The AHP-EIA was structured as shown if Fig 5. The questionnaire (shown in the S1 Appendix) that facilitated the performance analysis of CAs was constructed based on the 14 CAs and had a high level of detail based on the guidelines of the Brazilian Society of Cerebrovascular Diseases [49].

**Fig 5.**
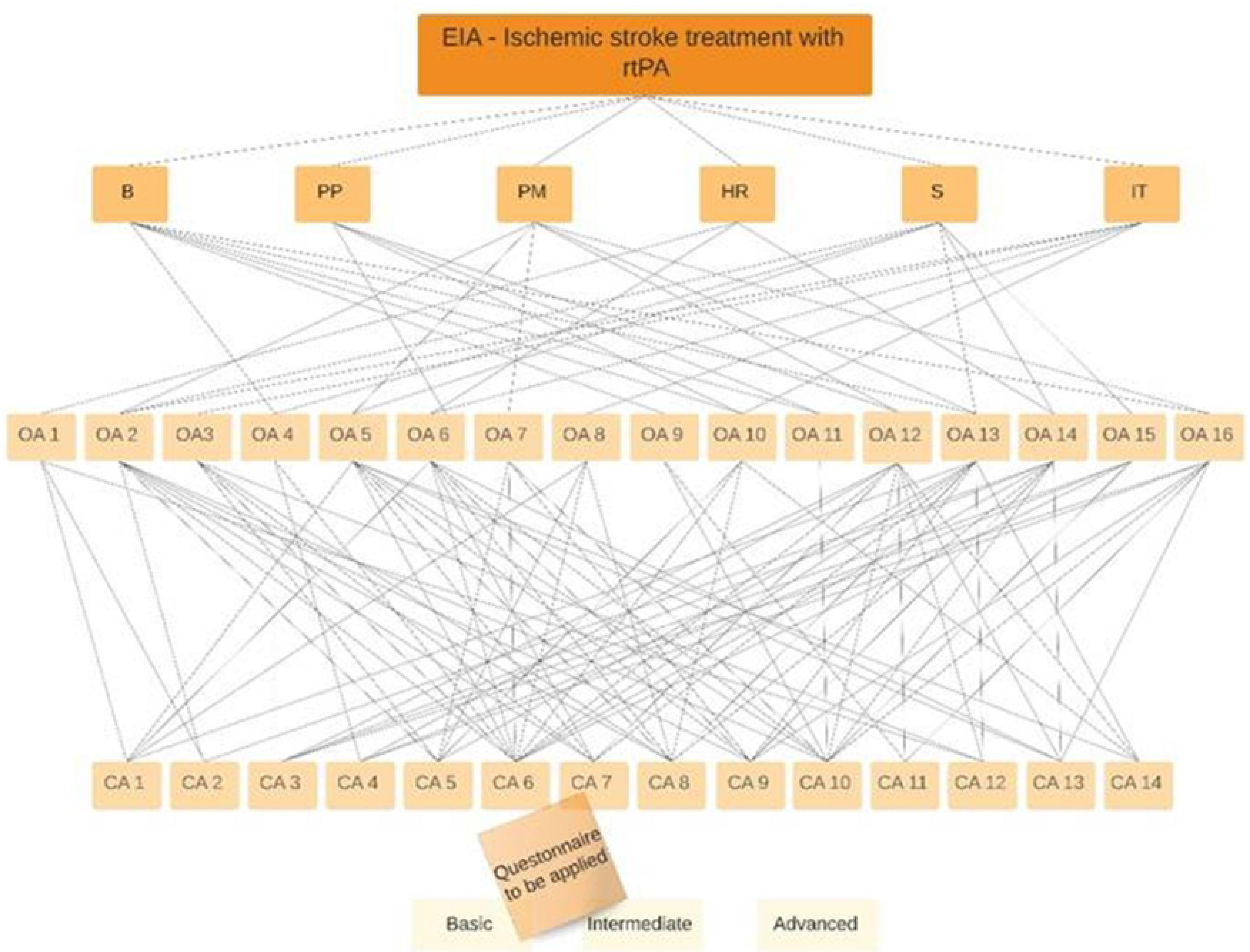
AHP structure designed for ischemic stroke process.

Facing a complex multi-criteria decision analysis, the AHP points out the part of the system that corresponds to each level of interoperability in the treatment of patients suffering from ischemic stroke with rtPA. This implies that there are concerns, regarding organizational and clinical attributes, that influence interoperability enhancement, others that demote it to a basic level, and still others that place it at the intermediate level. The results of the EIA, as shown in Fig 6, expressed that 33.36% were at the advanced level; 20.52% were at the basic level; 46.12% were at the intermediate level. This means that the UH was at an intermediate level of enterprise interoperability regarding ischemic stroke treatment.

**Fig 6.**
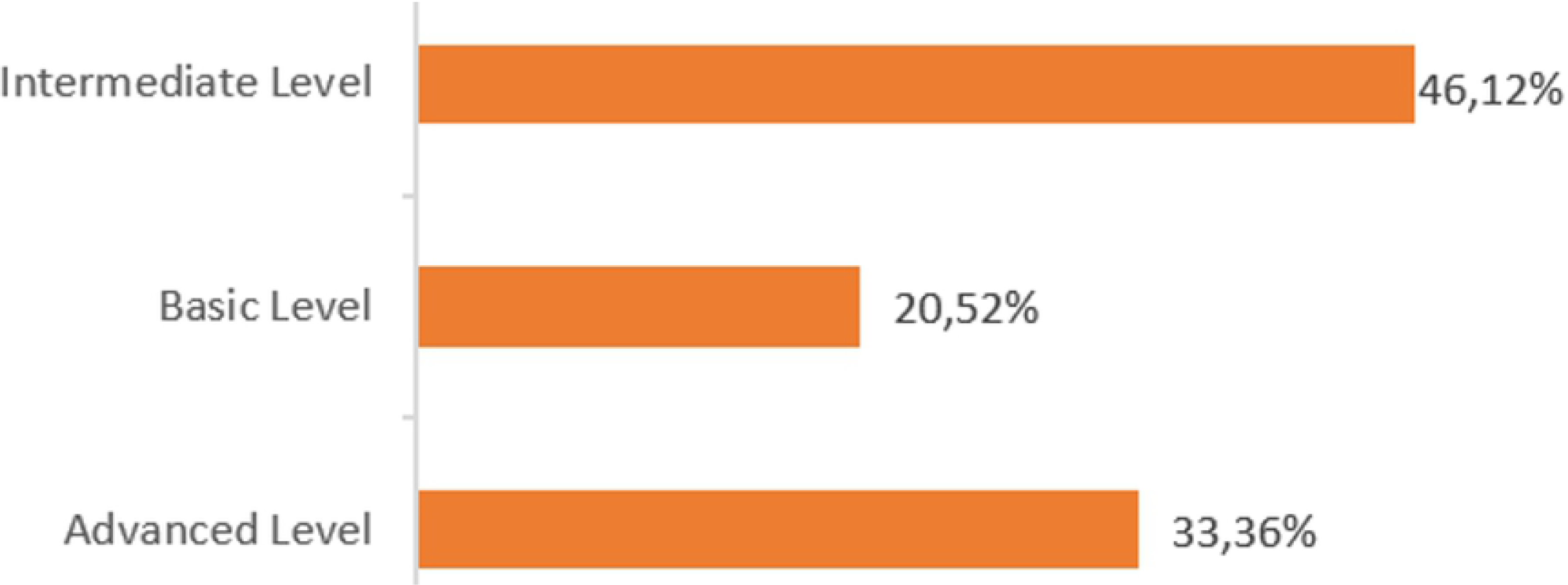
Result of the AHP-based EIA.

However, it should be noted that there was a relatively small difference between the intermediate level and the other two levels. This distributed diagnostic profile suggests performing a sensitivity analysis to understand how and when the current interoperability level could be superimposed by another. Analyzing sensitivity in decision-making methods targets the evaluation of the behavior of “alternatives” whenever there is a change in the “criteria” input data. Thus, in this case, it evidenced the influence that each CA had on the interoperability level.

Hence, sensitivity analyses were performed for each of the CAs, presented in Fig 7. The graphs demonstrate the behavior of the interoperability levels (lines) according to the prioritization given to each CA (x-axis). The extreme left of the x-axis is 0, a scenario in which no priority was given to such an attribute (so that it had an insignificant performance), and the opposite of the same axis represents the scenario in which a given attribute was given full priority with the maximum performance. The y-axis represents the level of enterprise interoperability reached.

**Fig 7.**
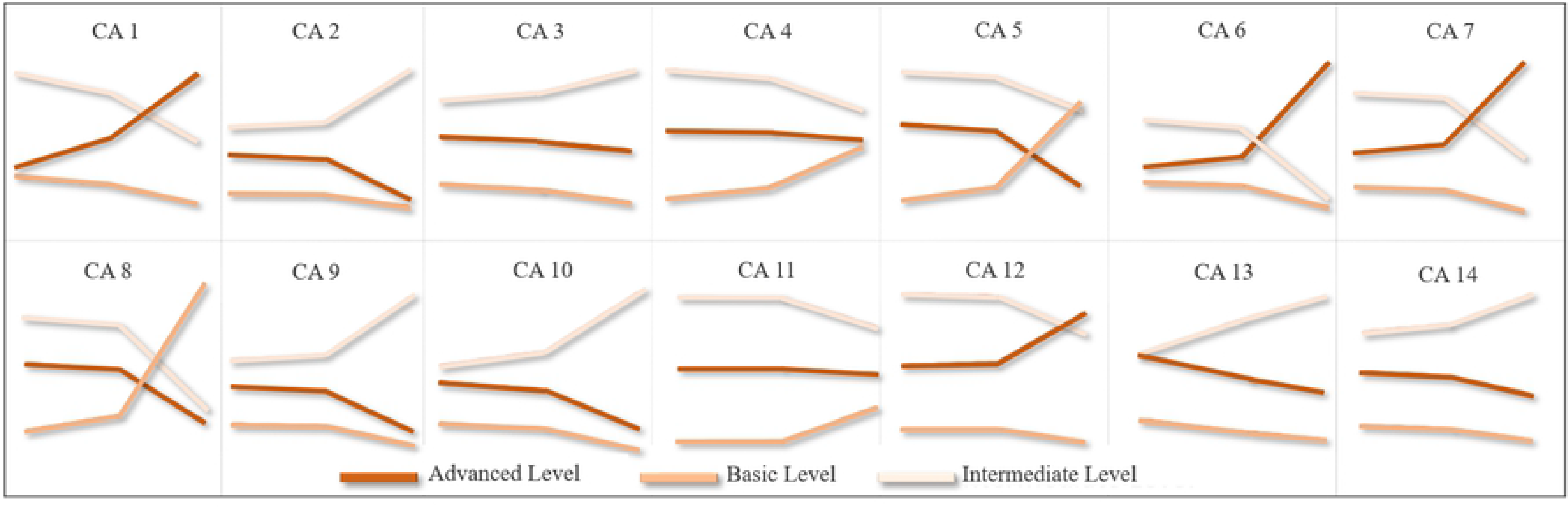
AHP sensitivity analysis. The darker curves represent the advanced level of enterprise interoperability, the medium-colored ones represent the intermediate level, and the lighter ones, the basic level.

Thus, for example, if the prioritization of Clinical Attribute 1 (CA 1 – Fig 8) is maximized, the attribute will leverage the global level of EIA (because the line that represents the advanced level will be in its upper half), but if its prioritization is minimized, the current overall EIA level (intermediate) will be maintained.

**Fig 8.**
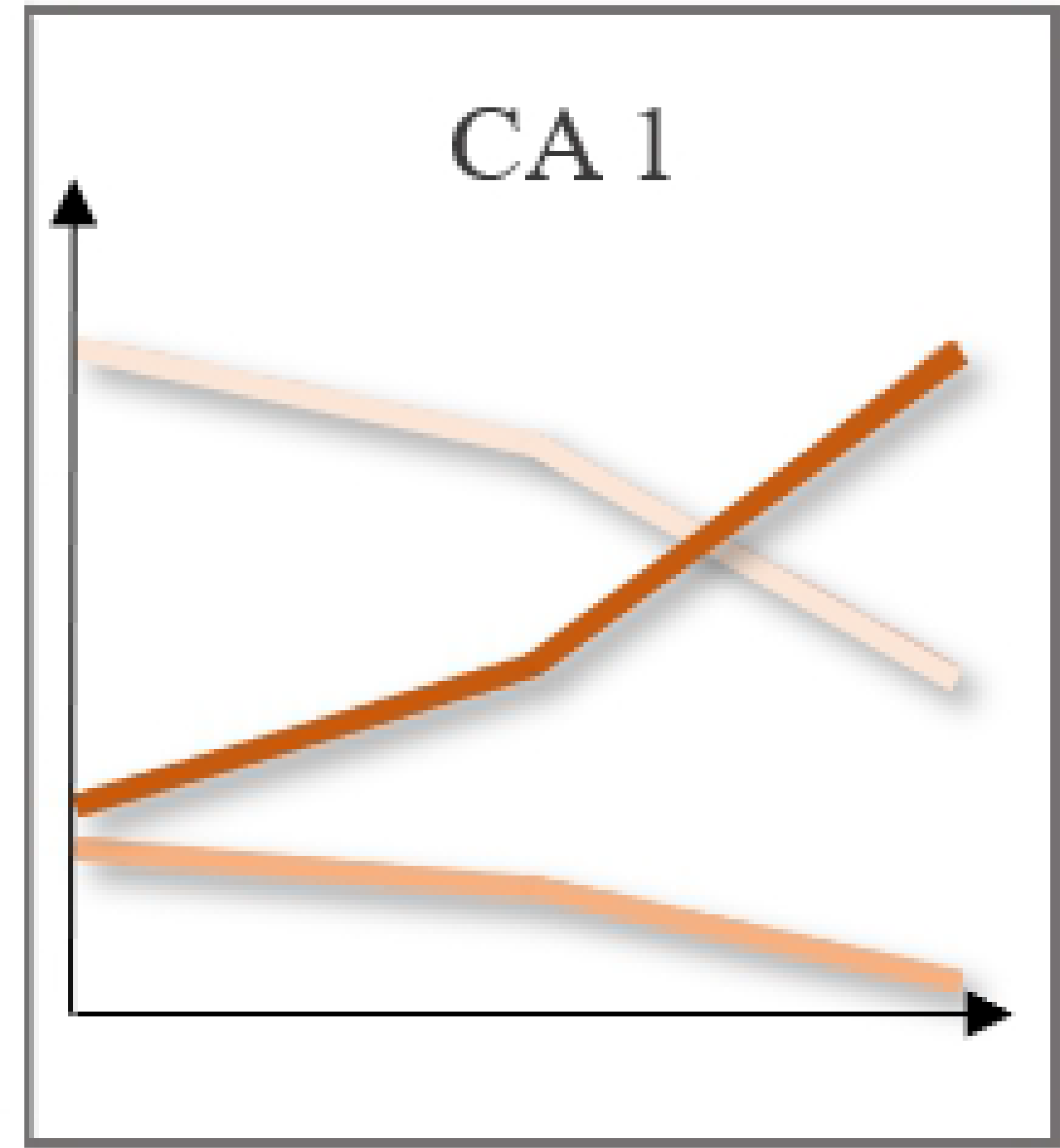
AHP sensitivity analysis (Clinical Attribute 1). Fig 9. AHP sensitivity analysis (Clinical Attribute 3).

On the other hand, if the Clinical Attribute 3 (CA 3 – Fig 9) receives the maximum or minimum of investment and efforts, even though more investments will tend to an advanced level, its EIA level will vary insignificantly. All the CAs can be analyzed following this rationale.

**Fig 9.**
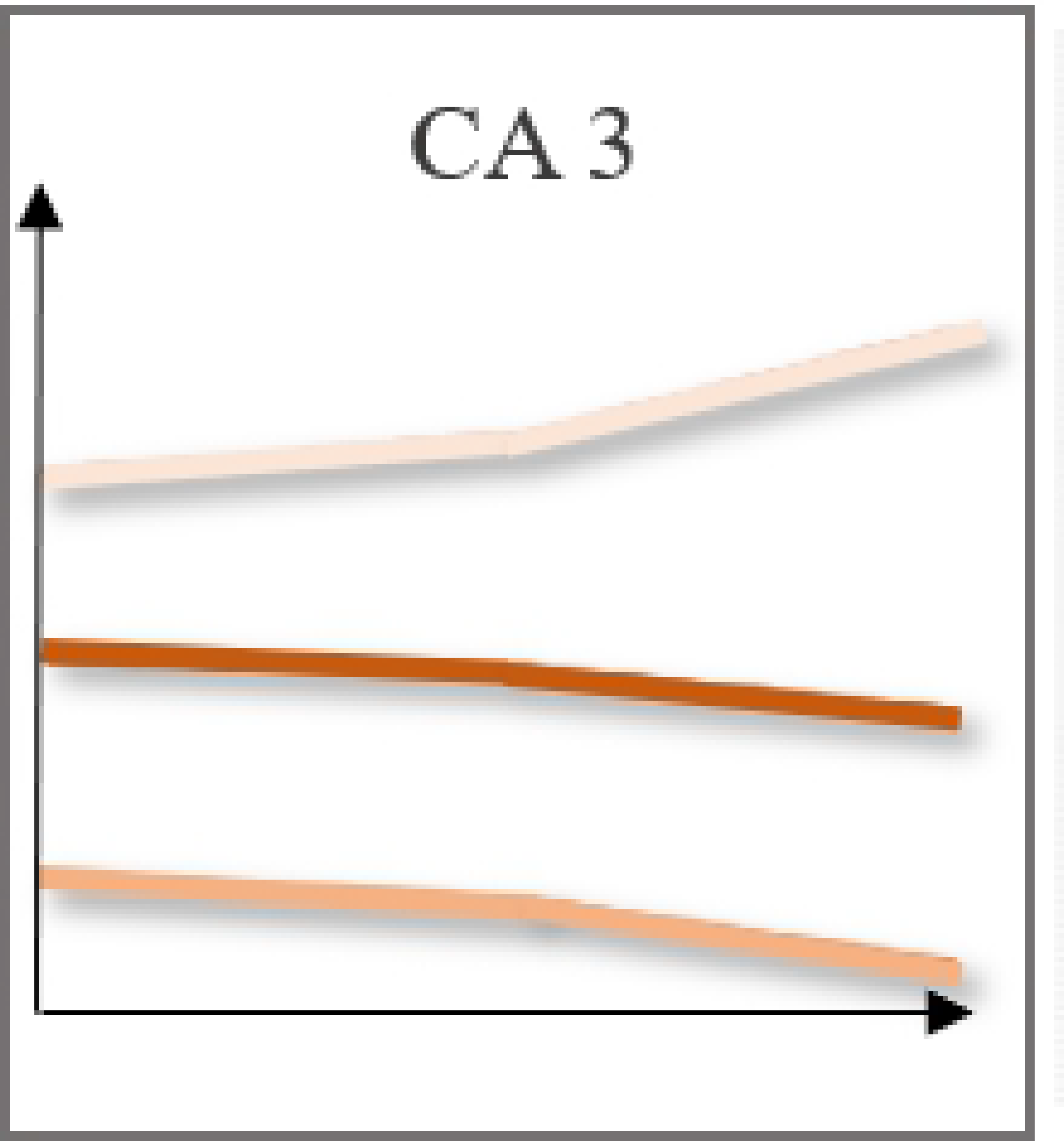
AHP sensitivity analysis (Clinical Attribute 3).

After the AHP sensitivity analysis was performed, it is notable that some CAs leveraged the global level of enterprise interoperability. These attributes are those that, the more prioritized they were, the more they tended to rise to the advanced level, and, at the same time, they tended to decrease to the basic and intermediate levels, as in cases: CA 1; CA 6; CA 7; CA 12.

### 2nd Stage: Development and PROMETHEE integration

This stage initiates with the calculation of the global weightings of the OAs (following Eq. 14), shown in Table 10.

**Table 10.** Global weightings of the OA.

| $W(AHP-concerns)$ | | $W(AHP-OA)$ | | | | | | | | | | | | | | | |
| --- | --- | --- | --- | --- | --- | --- | --- | --- | --- | --- | --- | --- | --- | --- | --- | --- | --- |
|  |  | OA 1 | OA 2 | OA 3 | OA 4 | OA 5 | OA 6 | OA 7 | OA 8 | OA 9 | OA 10 | OA 11 | OA 12 | OA 13 | OA 14 | OA 15 | OA 16 |
| (B) | 3,06 | 0 | 0 | 0 | 0,28 | 0 | 0 | 0 | 0 | 0,04 | 0,15 | 0,08 | 0 | 0,36 | 0 | 0 | 0,10 |
| (PP) | 11,98 | 0 | 0 | 0 | 0 | 0 | 0,36 | 0 | 0 | 0 | 0,11 | 0,05 | 0,48 | 0 | 0 | 0 | 0 |
| (PM) | 17,46 | 0 | 0,34 | 0 | 0 | 0,06 | 0 | 0,14 | 0 | 0 | 0 | 0 | 0,18 | 0,25 | 0 | 0 | 0,03 |
| (HR) | 38,65 | 0,24 | 0,31 | 0 | 0 | 0 | 0,14 | 0 | 0 | 0 | 0 | 0 | 0 | 0 | 0,31 | 0 | 0 |
| (S) | 25,40 | 0 | 0,25 | 0 | 0,07 | 0,05 | 0 | 0 | 0 | 0 | 0 | 0 | 0 | 0,14 | 0,26 | 0,22 | 0 |
| (IT) | 3,39 | 0 | 0 | 0,33 | 0 | 0 | 0,28 | 0 | 0,11 | 0,12 | 0,16 | 0 | 0 | 0 | 0 | 0 | 0 |
| $W(OA)$ | | | | | | | | | | | | | | | | | |
| Sum product function |  | 9,30 | 24,3 | 1,11 | 2,73 | 2,23 | 10,74 | 2,49 | 0,37 | 0,53 | 2,34 | 0,83 | 8,86 | 9,17 | 18,64 | 5,54 | 0,79 |
| Divided by 10 |  | 0,93 | 2,43 | 0,11 | 0,27 | 0,22 | 1,07 | 0,25 | 0,04 | 0,05 | 0,23 | 0,08 | 0,89 | 0,92 | 1,86 | 0,55 | 0,08 |

The AHP weightings of the concerns ( $w(AHP-concerns)$ ) are in the columns and AHP weightings of the OA ( $w(AHP-OA)$ ), in the rows. Their products are in the intersections. The global weightings of the OA ( $w(OA)$ ) are in the last row.

The OAs were organized in ascending order, enabling the inversion of the weightings from one MCDA to the other one. Thus, the OAs with the highest weightings in the AHP received the lowest weightings in the PROMETHEE II and so on. The PROMETHEE II criteria weightings were designated according to this rationale.

In addition to the criteria weights, the selected PROMETHEE preference functions were the usual and level ones, the most appropriate according to the behavior of the decision matrix. With Visual Promethee software, the values of *ϕ* were calculated for each concern in order to obtain the results exhibited in Table 11.

**Table 11.**
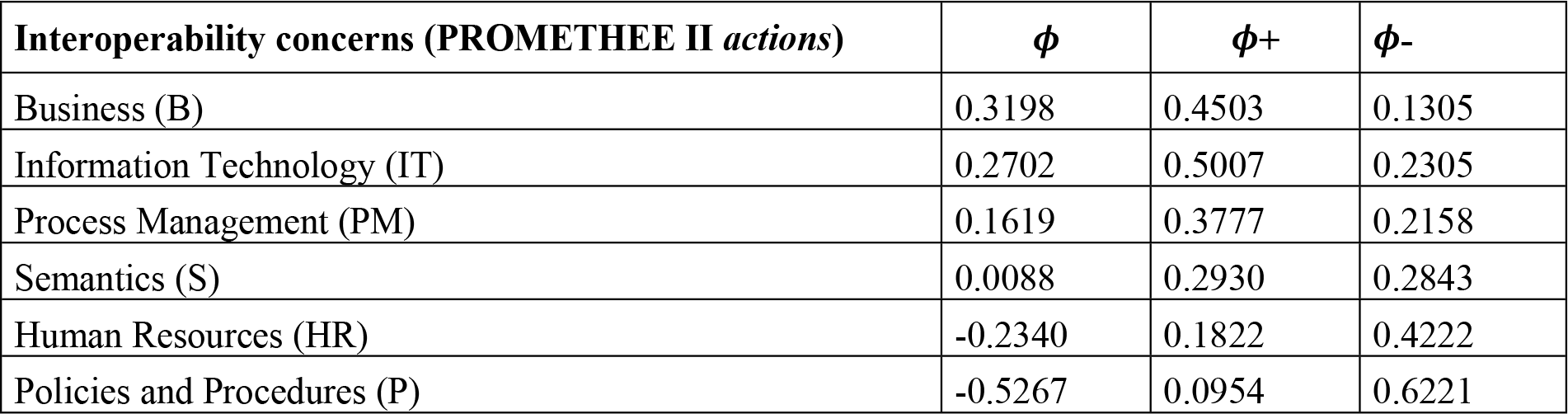
*ϕ* values of the Interoperability concerns

| Interoperability concerns (PROMETHEE II actions) | $\phi$ | $\phi+$ | $\phi-$ |
| --- | --- | --- | --- |
| Business (B) | 0.3198 | 0.4503 | 0.1305 |
| Information Technology (IT) | 0.2702 | 0.5007 | 0.2305 |
| Process Management (PM) | 0.1619 | 0.3777 | 0.2158 |
| Semantics (S) | 0.0088 | 0.2930 | 0.2843 |
| Human Resources (HR) | -0.2340 | 0.1822 | 0.4222 |
| Policies and Procedures (P) | -0.5267 | 0.0954 | 0.6221 |

Since *ϕ*+ expresses how much a given concern needs to be prioritized to overcome the others and *ϕ*- indicates by how much its performance is already sufficient to achieve the desired level of interoperability, it is possible to infer that those requiring more attention and efforts are Business and Information Technology, since they have a *ϕ* of 0.3198 and 0.2702, respectively. The concerns Policies and Procedures and Human Resources appear (*ϕ* -0.5267 and -0.2340, respectively) to be the most compliant with the requirements for a desirable level, that is, there is no need to change the OAs that represent them.

The three OAs with the highest standard deviation were chosen for a sensitivity analysis due to the direct relation between the deviation and the prioritization the hospital gives the OA. These were OA 2 – Electronic medical records, OA 12 – Compliance between practice and standards, and OA 13 – Coordination of clinical information flow.

A sensitivity analysis visualizes the impact made by an OA, that is, it allows for the verification of the behavior of the concerns when the organization changes its priorities regarding these OAs. Fig 10, utilizing the *ϕ* values, illustrates the current scenario, which shows that Human Resources and Policies and Procedures were already at a satisfactory level of interoperation and efforts for improvement must be invested in the other concerns. Fig 11 represents the sensitivity analysis that evaluated OA 2, OA 12, and OA 13.

**Fig 10.**
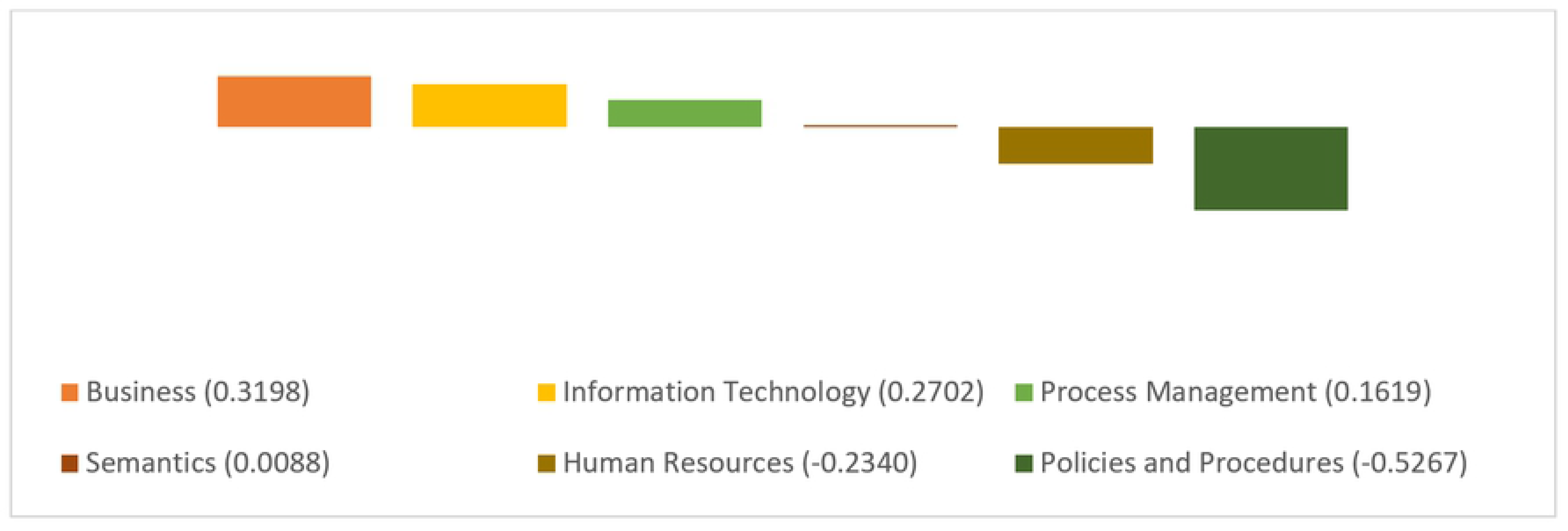
Current scenario of Interoperability concerns.

**Fig 11.**
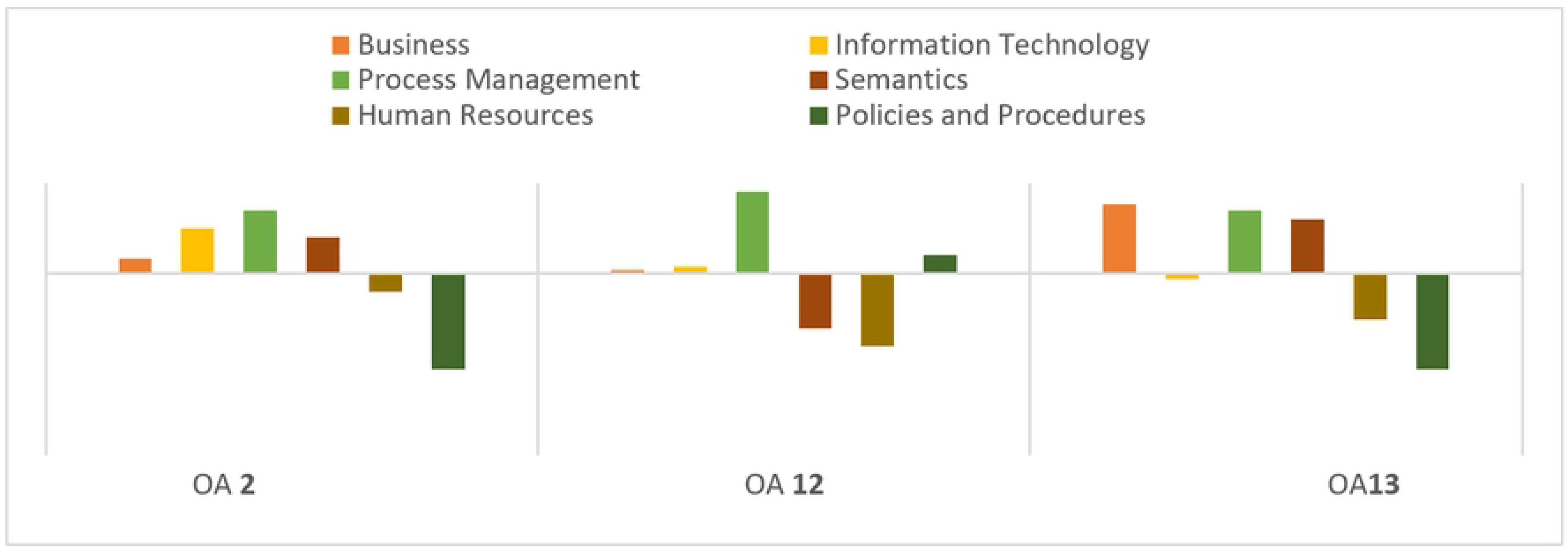
Sensitivity analysis of the organizational attributes 2 (left), 12 (middle) and 13 (right).

In these three sensitivity analyses, the prioritization values were changed until a “turning point” was found, i.e., a percentage of prioritization of the OA that resulted in a significant change in the concerns. The first attribute analyzed was OA 2 - Electronic Medical Record, whose priority was changed from 0 to 25%, the “turning point.” This change in OA 2 impacted the Process Management (PM) concern, turning it into the new strategic focus.

The second attribute analyzed was OA 12 *-* Compliance between practice and standards. Its weighting was increased from 2% to 47%, with the others decreasing proportionally. This prioritization generated expressive modifications in all interoperability concerns. In Business (B), this determined whether the flow would be positive or negative. If it kept increasing, the flow would become negative. In Information Technology (IT), the flow became negative, expressing an improvement in this concern if OA 12 was prioritized. Process Management (PM) continued to be positive. Semantics (S) and Human Resources (HR) were in the negative flow. Policy and Procedures (P) crossed the axis of *ϕ*.

Finally, the analysis of OA 13 - Coordination of clinical information flows - changed OA 13’s prioritization from 1% to 66%. The most significant change that could be perceived was related to a better performance recorded by the Information Technology (IT) concern.

## Discussion of the results of the case study

The 1^st^ stage of this framework enabled the UH’s EIA, positioning this hospital at the intermediate level of interoperability concerning acute ischemic stroke treatment. This means that, as per the healthcare interoperability levels proposed, the UH is capable of assisting patients with stroke symptoms and striving to decrease its process variability levels and increase the integration among its sectors. As per the proposed classification, its complex systems and processes are centralized by a single entity (information system), achieving a certain degree of integration among different stakeholder entities. This practice is reflected in the result since, at the time of this study, the UH was in a transitional phase, moving toward a unified information system. However, there are various areas related to interoperation that need to be optimized.

As the basic and advanced level components were meaningful in this result, a sensitivity analysis of CAs was carried out in this first stage. This indicated that four CAs tended to leverage enterprise interoperability. These CAs were CA 1 - Initial measurement of vital signs; CA 6 - Differential diagnosis; CA 7 - Neurological status; and CA 12 - Establishing the exact time of symptoms onset.

The S2 pointed out that the interoperability concerns that needed more prioritization were Business (B) and Information Technology (IT). As previously defined, Business (B) enhances enterprise interoperability from the point of view of strategic aspects of the organization and its relations with external and internal entities. Information Technology (IT) includes technologies and information systems that help to improve and optimize processes. The 2^nd^ Stage also showed that the OAs that most impacted these concerns were OA 2 - Electronic medical records, OA 12 - Compliance between practice and standards, and OA 13 - Coordination of clinical information flows. This suggests that UH managers and staff should dedicate more effort to these attributes, especially from the purview of Information Technology and Business. This could be translated as the need to (i) invest more in an easier and feasible information system, with data security and availability; (ii) update medical equipment, tools, and technological resources involved in the process according to the best and latest clinical practices and guidelines; (iii) align technology and its maintenance with the hospital’s organizational strategy; and (iv) have effective intra and interorganizational integration among different sectors, hierarchical levels, and information systems.

## Conclusion and future research

This research proposed a framework that structured an EIA to optimize time-dependent and critical clinical processes. Initially, a literature review was conducted to define the concepts of interoperability and its frameworks in the context of HC, to understand the ED’s processes and comprehend the usage of the MCDA methods, the AHP and PROMETHEE II. Supported by the literature, the use of the hybrid approach between the AHP and PROMETHEE II was assertive, due to the effective complementarity between both. Furthermore, the understanding of the process was consolidated based on interviews with experts. Thereafter, the attributes related to these processes were listed and classified as either organizational or clinical.

After all, the proposed framework was implemented during a case study in a UH in the south of Brazil and, thus, validated. The framework proved to be able to diagnose the interoperability level of the UH’s processes as well as point out how this level could be changed. Furthermore, the diagnostical perspective could be optimized if the decision making trial and evaluation laboratory (DEMATEL), another MCDA, was employed. Aggregating this analysis is suggested as future research since the MCDA approach identifies the cause-effect chain between elements.

It is also important to note that the design of the proposed framework can be adapted (in relation to CAs) and applied to other processes in the HC domain. Thus, for future works, it is also suggested that such assessments are carried out in other hospital departments as well. The main obstacle faced during this research was the initial restriction of communication with the physicians involved in the case study. They were initially retiscent until they comprehended the practical contribution of this research to their medical routine.

## Abbreviations

ED: Emergency Department
MCDA: multicriteria decision analysis methods
AHP: Analytic Hierarchy Process
PROMETHEE: Preference Ranking Organization Method for Enrichment Evaluation
EIA: Enterprise Interoperability Assessment
CA: clinical attributes
HC: healthcare
OA: organizational attributes
UH: University Hospital.

## Data Availability

All relevant data are within the manuscript and its Supporting Information files

## Supporting Information

S1 Appendix. The questionnaire that allowed the performance analysis of CAs.

